# Activation of the vagal anti-inflammatory reflex by remote ischaemic conditioning in humans: experimental cross-over study

**DOI:** 10.1101/2021.01.28.21249488

**Authors:** Shaun M. May, Eric Chiang, Anna Reyes, Gladys Martir, Amour Patel, Shamir Karmali, Sanjiv Patel, Simeon West, Ana Gutierrez del Arroyo, Alexander V. Gourine, Gareth L. Ackland

## Abstract

**BACKGROUND:** Non-invasive approaches in humans that may activate the vagal anti-inflammatory reflex are lacking. Neurons within the dorsal motor vagal nucleus (DMVN) activate both the vagal anti-inflammatory reflex (which regulates leukocyte trafficking by controlling neutrophil surface CD11b expression) and cardioprotection afforded by remote ischemic conditioning (RIC). We tested the hypothesis that RIC recruits vagal activity and activates the anti-inflammatory reflex in humans by reducing neutrophil (CD16^+^)CD11b expression.

**METHODS:** Participants (age:50±19 years; 53% female) underwent ultrasound-guided injection of local anaesthetic within the brachial plexus before applying 37×8 min cycles of brachial artery occlusion using a blood pressure cuff (RIC_block_). RIC was repeated 6 weeks later without brachial plexus block. Masked analysers quantified vagal activity (heart rate variability) before, and 10 minutes after, the last RIC cycle. The primary outcome was RR-interval, compared between RIC_block_ and RIC. Secondary outcomes were time-domain, frequency-domain, and flow cytometric quantification of CD16^+^CD11b expression in whole blood (incubated with lipopolysaccharide (LPS) or saline) compared between RIC_block_ and RIC.

**RESULTS:** RIC increased RR-interval (lowered heart rate) by 40ms (95% confidence intervals (95%CI):13-66; n=17; *P*=0.003). RR-interval did not change after RIC_block_ (mean difference:20ms (95%CI:-11 to 50); *P*=0.19). High-frequency (vagal) modulation of heart rate was reduced after RIC_block_, but preserved after RIC (*P*<0.001). indicating RIC preserved vagal activity. LPS-induced CD16^+^CD11b^+^ expression was lower after RIC (3615 median fluorescence units (95%CI:475-6754); *P=*0.026), compared with 2331 units (95%CI:-3921 to 8582); *P=*0.726) after RIC_block_.

**CONCLUSION:** RIC recruits the vagal anti-inflammatory reflex, which requires intact afferent signalling from the peripheral tissue undergoing ischaemia/reperfusion to increase vagal tone and reduce neutrophil activation.

**TRIAL REGISTRATION:** researchregistry6482.

---

Electronic devices that stimulate the vagus nerve inhibit inflammation, cytokine production and neutrophil CD11b surface expression through the cholinergic anti-inflammatory pathway.(1) The key neural substrate for this neuro-immune pathway originates from cholinergic neurons residing within the brainstem dorsal vagal motor nucleus (DVMN) that project to the celiac-superior mesenteric ganglia to increase splenic nerve activity and inhibit inflammation.(2) Acetylcholine signalling through α7 nicotinic acetylcholine receptors (α7nAChR) inhibits release of TNFα from splenic macrophages and suppresses F-actin polymerization, the rate-limiting step for CD11b surface expression on circulating neutrophils.(3) CD11b is a critical β_2_-integrin regulating neutrophil adhesion to the endothelium and transmigration to sites of injury and infection.(4) Modulation of leukocyte trafficking via cholinergic signaling thus suppresses the excessive accumulation of neutrophils at inflammatory sites.(3)

DVMN neurons are also critical for remote ischaemic conditioning (RIC), (5, 6) which preserves cardiac tissue(7) and exercise capacity after myocardial infarction in rats(8) through time-limited, repetitive ischaemia-reperfusion in a distant limb.(9, 10) Chemogenetic silencing of vagal neurons prevents RIC from reducing the extent of injury after myocardial infarction.(7) Since RIC and the cholinergic anti-inflammatory pathway are both cardioprotective, (11) they appaear likely to share common neural substrate residing within DVMN.

Whilst experimental activation of the inflammatory reflex has been studied using specific vagus nerve-stimulating devices, non-invasive approaches to activate the vagal anti-inflammatory reflex in humans that can be applied at scale are lacking. Laboratory data demonstrate that RIC requires intact afferent sensory innervation of the remote organ undergoing RIC.(12, 13) Therefore, we exploited this neurophysiologic feature to test the hypothesis that nerve blockade at the level of the brachial plexus prevents cardiac vagal and/or anti-inflammatory effects of RIC by interrupting the afferent arm of the reflex in humans. We provide the first proof-of-concept data in humans showing that RIC recruits the vagal anti-inflammatory reflex.

## Methods

### Study design

We conducted an experimental cross-over study in subjects scheduled for upper limb surgery at University College London Hospitals NHS Trust. The study was conducted in accordance with the principles of the Declaration of Helsinki and approved by the London (Stanmore) Research Ethics Committee (16/LO/0634). Subjects provided written informed consent to participate, having given verbal consent at least 72h before the first intervention.

### Inclusion/exclusion criteria

Subjects aged >18y scheduled for elective upper limb surgery requiring regional anaesthesia alone were eligible. Exclusion criteria were the presence of upper limb pathology precluding use of blood pressure cuff, previous splenectomy, a history of cardiac arrythmias and/or abnormal preoperative electrocardiogram (conduction abnormality-bundle branch block, bradyarrhythmia), allergy to local anaesthetic agents and failure to achieve loss of motor power after ultrasound guided delivery of local anaesthetic.

### Protocol

Subjects first underwent RIC after brachial plexus block was established, prior to upper limb surgery (RIC_block_ in text; Figure 1A). On return to hospital for outpatient follow-up at least 5 weeks after surgery, RIC was repeated in the absence of brachial plexus analgesia over the same timeframe as for the first visit (Figure 1B).

**Figure 1.**
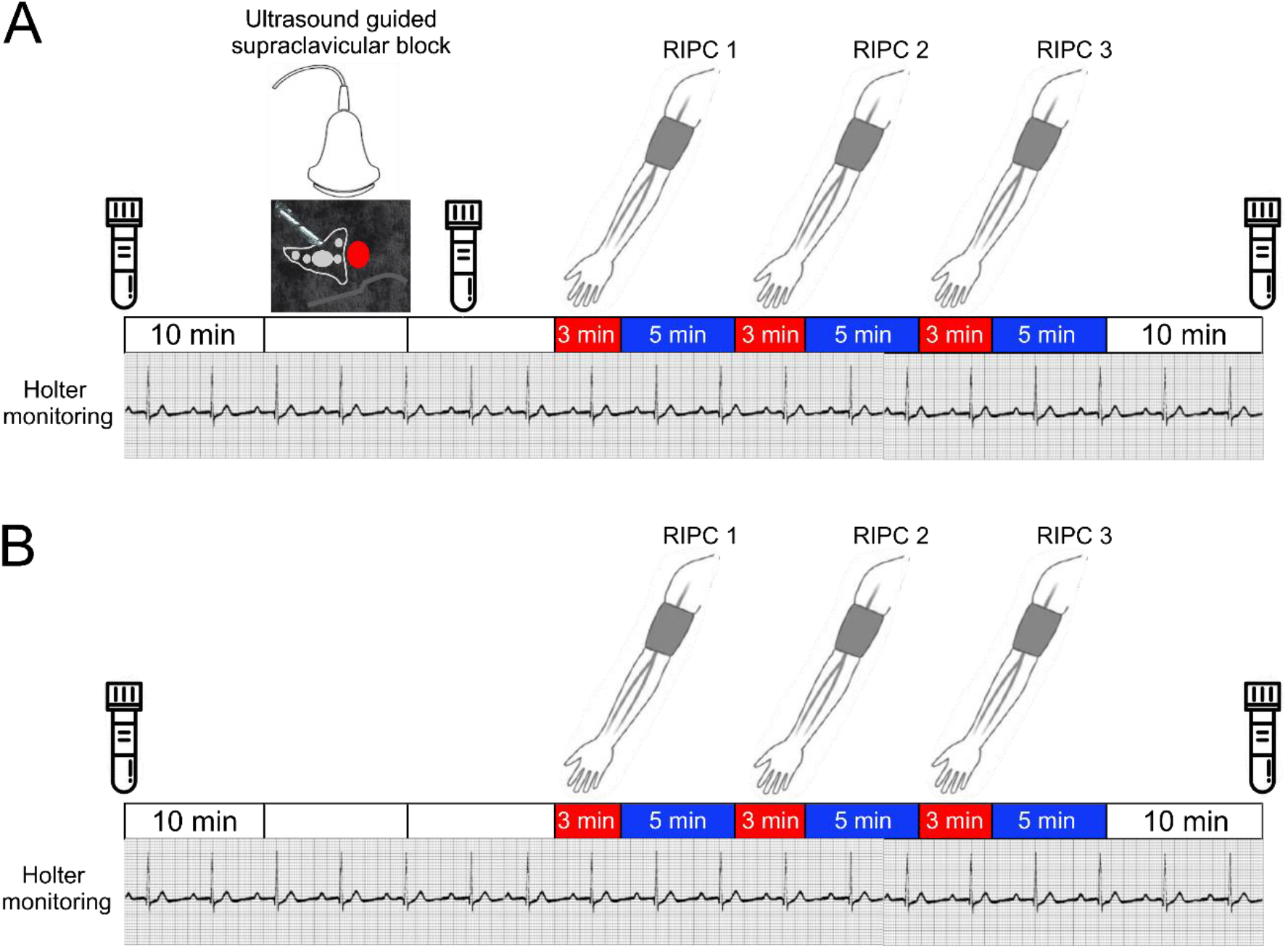
Experimental design. Sequence of experimental procedures for RIC undertaken in presence and absence of brachial plexus block. 10 minute recording sessions are indicated by clear bars. Three cycles of ischaemia (applied for 3 minutes) followed by 5 minutes of reperfusion were applied. Blood samples were obtained at the times indicated by the collection bottles. A. For the first visit, brachial plexus block with ultrasound guided injection of local anaesthetic was undertaken. B. For the return visit, a time-matched protocol was followed without brachial plexus block before three cycles of RIC were undertaken.

### Ultrasound-guided supraclavicular brachial plexus block

All ultrasound-guided blocks were conducted by senior anaesthetic staff before surgery in a quiet dedicated procedure room (Figure 1A). Patients were positioned in a semi-sitting position with the head turned 45 degrees to the non-operative side. Standard cardiorespiratory monitoring plus Holter monitors were applied. Peripheral intravenous access was established but no sedation was administered. Under sterile conditions, a high-frequency linear array transducer (13–6 MHz, SonoSite S-Nerve; SonoSite, Inc., Bothell, WA, USA) probe was placed over the supraclavicular fossa, parallel to the clavicle to obtain a short-axis view of the divisions of the brachial plexus and the subclavian artery, lying on the first rib. Following skin infiltration with lidocaine 1%, a 22-gauge 50 mm insulated echogenic needle (Pajunk, Geisingen, Germany) was guided in-plane with the ultrasound beam until the needle tip was positioned at the junction of the first rib and subclavian artery; 20ml 0.5% bupivacaine was injected using a single-injection technique, with intermittent aspiration under constant ultrasound visualisation. If paraesthesia developed, the needle tip was repositioned. After completion of the block, patients remained fully monitored until their transfer to the operating room. Assessment of sensory and motor blocks was performed by one of the investigators (SK, EC, SM, GM, AR) every 5 min after local anaesthetic injection. Inability of the anaesthetised arm to overcome gravity was required prior to beginning RIC.

### Upper limb remote ischaemic preconditioning

Three cycles of ischaemia/reperfusion were performed, by inflating a 12cm wide blood pressure cuff over the upper arm for 3 minutes above the systolic blood pressure to occlude the brachial artery before cuff deflation for 5 minutes (Figure 1).

### Heart rate analysis

Three lead electrocardiogram recordings were captured using Lifecard CF digital Holter monitors (Spacelabs Healthcare, Hertford, UK), with a sampling rate of 1024Hz. R-R intervals from ECG data were analysed after data cleaning. Heart rate variability (HRV) analysis from the final 5 minute segments within each recording periods by analysers masked to the intervention (Figure 1), in accord with Taskforce guidelines.(14) Serial changes in cardiac autonomic activity were quantified using three established measures of autonomic modulation of heart rate; time domain, frequency domain and non-linear analyses (Kubios HRV Premium software, Version 3.1.0, Kuopio, Finland)(15), as described previously.(16) For time-domain measures, we analysed RR interval, minimum and maximum heart rates. We performed spectral analysis using the parametric autoregressive method (AR) because it produces a spectrum with better resolution when short data frames are used and the spectrum can be divided into independent components.(17) We used high frequency (0.15-0.4 Hz) power spectral analysis of R-R interval time series (AR spectrum), as a measure of cardiac vagal activity.(14) Low frequency (0.04-0.15Hz) power spectral analysis was also assessed as a measure of arterial baroreflex sensitivity.(18) We also examined detrended fluctuation non-linear analysis correlation measures, in which a series of RR intervals are integrated and are divided into a series of regular intervals. For each interval the fluctuation of the data from a straight line of linear interpolation is calculated. We examined changes in DFAα1, the values of which increase with pharmacologic (atropine) vagal blockade and decrease with sympathetic blockade.(19)

### Flow cytometry

Whole blood was collected from patients who consented to repeat blood draws in BD Vacutainer® sodium citrate tubes (Becton Dickinson, UK) before the first ten-minute period of heart rate recording and at the end of the final ten-minute period of heart rate recording. To determine whether vagal activation by RIC may modulate acute inflammation, we examined whether the presence – or absence-of brachial plexus block altered myeloid (neutrophil, monocyte) cell activation after the incubation with lipopolysaccharide (LPS). Samples were incubated with, or without, low dose of LPS (10 ηg ml^-1^; *Escherichia coli* endotoxin, serotype O111:*B4*, Sigma) at 37°C for 2 hours with gentle agitation. Immediately before the start of the protocol, and 10 minutes after the final RIC cycle, 100µl whole blood samples were immediately stained as described in Supplementary data (including gating strategy and antibodies used). Analysis was performed by investigators masked to the intervention. Neutrophils and monocytes were identified using forward and side scatter characteristics (Supplementary figure 1), in combination with specific cell surface antigen for CD16 (clone VEP13), CD14 (clone TUK4), respectively (Miltenyi Biotec, Germany). Co-expression of surface CD11b, CD182 [CXCR2], HLA-DR (Miltenyi Biotec, Germany) were quantified, using frequency-minus-one and appropriate isotype controls. Acquired data (BD LSRFortessa) were analyzed using FlowJo (BD Biosciences, Oxford, UK) software.

### Statistical Analysis

Manual and automated validation checks of data were undertaken. Categorical data are summarised as absolute values (percentage). Continuous data are presented as mean (SD), or median [IQR], unless stated otherwise. Repeated-measures analysis of variance was used to compare heart rate and heart rate variability measures before and after RIC, taking into account each subject, timepoint and intervention (RIC with/without nerve block). Individual comparisons between groups were calculated using post-hoc Tukey-Kramer tests. Statistical analyses were undertaken using NCSS 2020 (Kaysville, UT, USA).

### Sample size calculation

With a predicted resting heart rate 80±10bpm, we estimated that a 10bpm difference (SD:10bpm) between RIC with versus without upper limb block would require at least 15 individuals to be recruited (α=0.01; 1-β=0.9). Allowing for 25% drop-out rate (including failure to re-attend for the follow-up protocol), the estimated sample size was initially increased to 30 subjects but revised to 20 subjects having established >90% follow-up rate from the first 10 participants.

## Results

### Subject characteristics

A minority of the participants were taking any regular cardiovascular medications. None had a clinical history of ischaemic heart disease. The single subject requiring oral medication for diabetic control did not have any clinical evidence for pre-existing neuropathy (Table 1).

**Table 1.** Subject characteristics. Data is presented as mean with standard deviations (SD) for parametric data and as median (25th-75th interquartile range) for non-parametric data. Frequencies are presented with percentages (%). Age is rounded to the nearest year. ACE-I: Angiotensin converting enzyme inhibitor. ARB: angiotensin receptor blocker.

|  |  |
| --- | --- |
| Age (y) | 56 (36-61) |
| Female sex (n;%) | 9 (52.9%) |
| **Co-morbidities (n; %)** |  |
| Asthma | 5 (29.4%) |
| Atherosclerotic cardiovascular disease | 0 |
| Diabetes mellitus | 1 (5.9%) |
| Current smoker | 4 (23.5%) |
| Hypertension | 2 (11.8%) |
| Active cancer | 0 (56.1%) |
| Inflammatory arthritis | 4 (23.5%) |
| **At least one cardiovascular medication (n; %)** | 4 (23.5%) |
| Beta blocker | 1 (5.9%) |
| Calcium channel blocker | 1 (5.9%) |
| Diuretic | 1 (5.9%) |
| Statin | 1 (5.9%) |
| ARB/ACE-I | 0 |
| Nitrate | 1 (5.9%) |
| NSAIDs | 2 (11.8%) |
| Opioids | 1 (5.9%) |
| Antidepressant | 2 (11.8%) |
| Steroid/immunosuppressant | 0 |

Most subjects were undergoing elective upper limb surgery for long-standing orthopaedic indications, rather than inflammatory arthropathy. Repeat blood samples were obtained from nine patients who consented to repeat sampling.

### Time domain measures of heart rate variability

RR-interval did not change after RIC_block_ (mean difference:20ms (95%CI:-11 to 50), P=0.19). RIC increased RR-interval (i.e. slowed heart rate) by 40ms (95% confidence intervals (95%CI):13-66). (Figure 3A). In the presence of brachial plexus block, minimum RR-interval after RIC did not change (mean difference:26ms (95%CI:-20 to 72); P=0.24) (Figure 3B). RIC increased minimum RR-interval by 39ms (95% CI:3-74); P=0.03). Similarly, after RIC_block_, maximum RR-interval remained unchanged (mean difference:12ms (−20 to 41); P=0.43; Figure 3C). Maximum RR-interval increased after RIC by 28ms (95% CI:-2 to 58); P=0.06). Other time-domain measures of HRV were similar between RIC and RIC_block_ (Supplementary figure 2).

**Figure 2.**
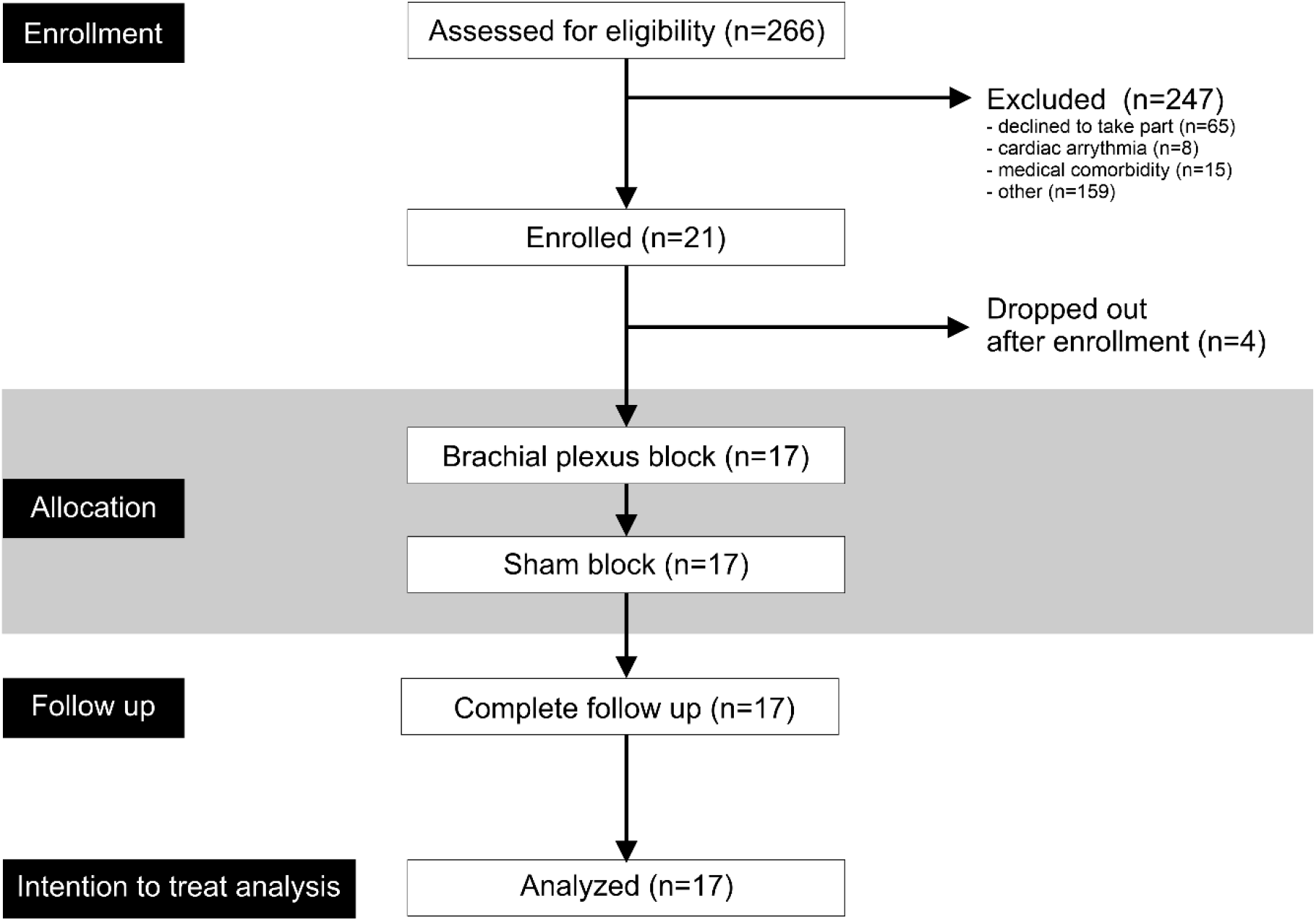
CONSORT diagram. Screening, enrolment and follow up data for study, in accord with CONSORT requirements.

**Figure 3.**
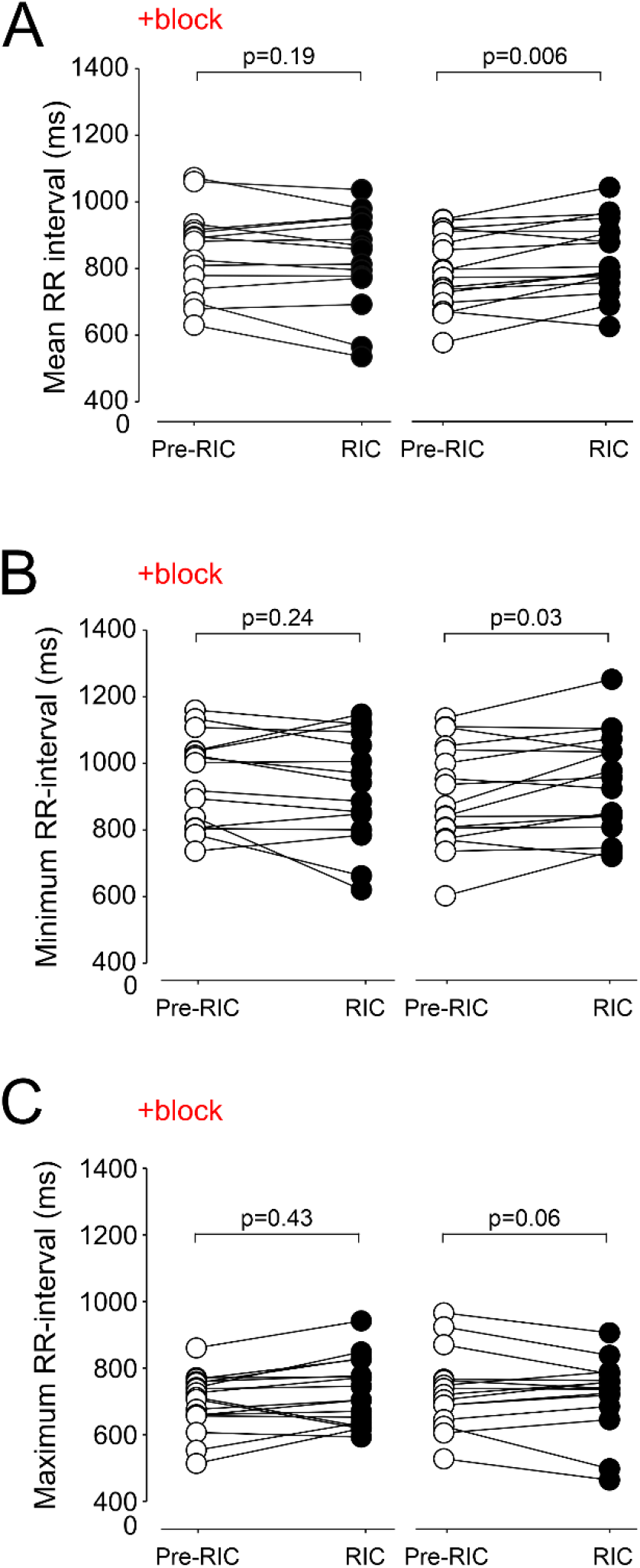
Heart rate changes before and after RIC in same individuals with/without brachial plexus block. Pre-data point for each experiment refers to final 5 minutes of data recorded within 10 minute period before brachial plexus block was performed. Post refers to final 5 minutes of data recorded within 10 minute time period following the final 5 minute washout period of the third cycle of RIC. A. Individual datapoints for mean R-R interval in the presence, and absence, of brachial plexus block, during pre and post-RIC recording periods. B. Individual datapoints for minimum heart rate achieved in the presence, and absence, of brachial plexus block, during pre and post-RIC recording periods., p values comparing pre versus post values (repeated-measures analysis of variance), post-hoc Tukey-Kramer test. C. Individual datapoints for maximum heart rate achieved in the presence, and absence, of brachial plexus block, during pre and post-RIC recording periods. p values comparing pre versus post values (repeated-measures analysis of variance), post-hoc Tukey-Kramer test.

### Frequency-domain measures of heart rate variability

Low frequency (LF) band peak frequency (autoregressive spectrum) did not change after RIC, but in the presence of brachial plexus block RIC resulted in increased LF component of HRV (Figure 4A). In the absence of neural block, RIC had no effect on the high-frequency (HF) vagal modulatory component of HRV (Figure 4B). In the presence of brachial plexus block, HF was reduced (intervention x timepoint: *F*_(3,3.26)_ P=0.03).

**Figure 4.**
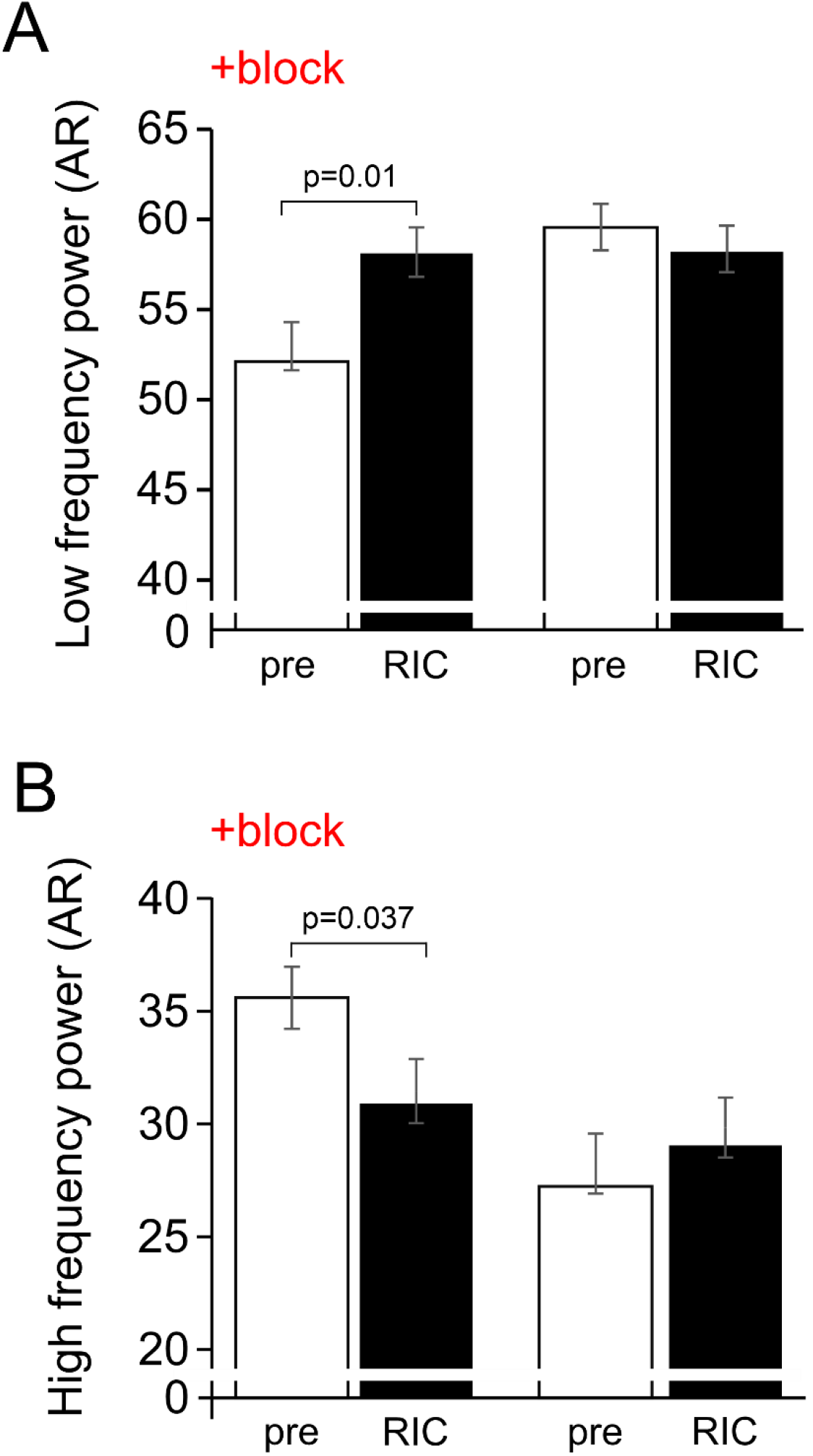
Frequency domain measures before and after RIC in same individuals with/without brachial plexus block. A. Low frequency power. B. High frequency power. Mean (95% confidence intervals) shown; values are shown for post-hoc Tukey-Kramer testing, following repeat-measures ANOVA.

### Non-linear analysis of heart rate variability

The detrended fluctuation analysis measure DFAα1 increased after RIC_block_ (0.124 (95% confidence intervals:0.01-0.24); P=0.039), indicative of reduced vagal tone. In the absence of brachial plexus block, DFAα1 remained unchanged after RIC (Figure 5).

**Figure 5.**
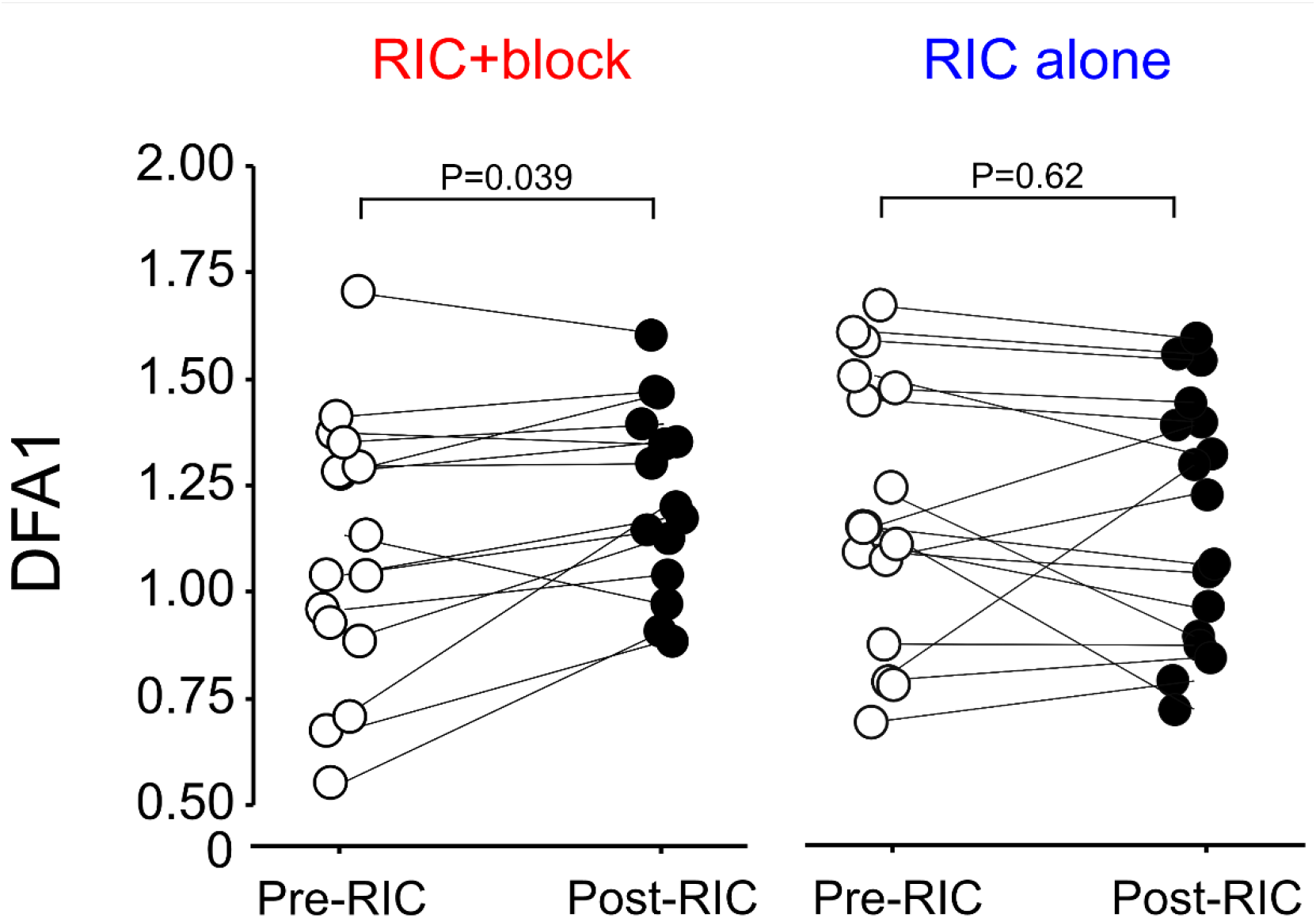
Non-linear analysis of HRV before and after RIC in same individuals with/without brachial plexus block. Individual datapoints for DFA in the presence, and absence, of brachial plexus block, pre and post RIC. p values refer to posthoc Tukey-Kramer testing, following repeat-measures ANOVA.

### Neutrophil activation following RIC

Repeat blood samples were obtained from nine patients who consented to repeat sampling. Before establishing brachial plexus blockade (Figure 6A), expression of CD11b on neutrophils increased after incubation with LPS in whole blood (mean difference in median fluorescence intensity (MFI): 19 (95%CI:9-28); P=0.001; post-hoc Tukey-Kramer test).

**Figure 6.**
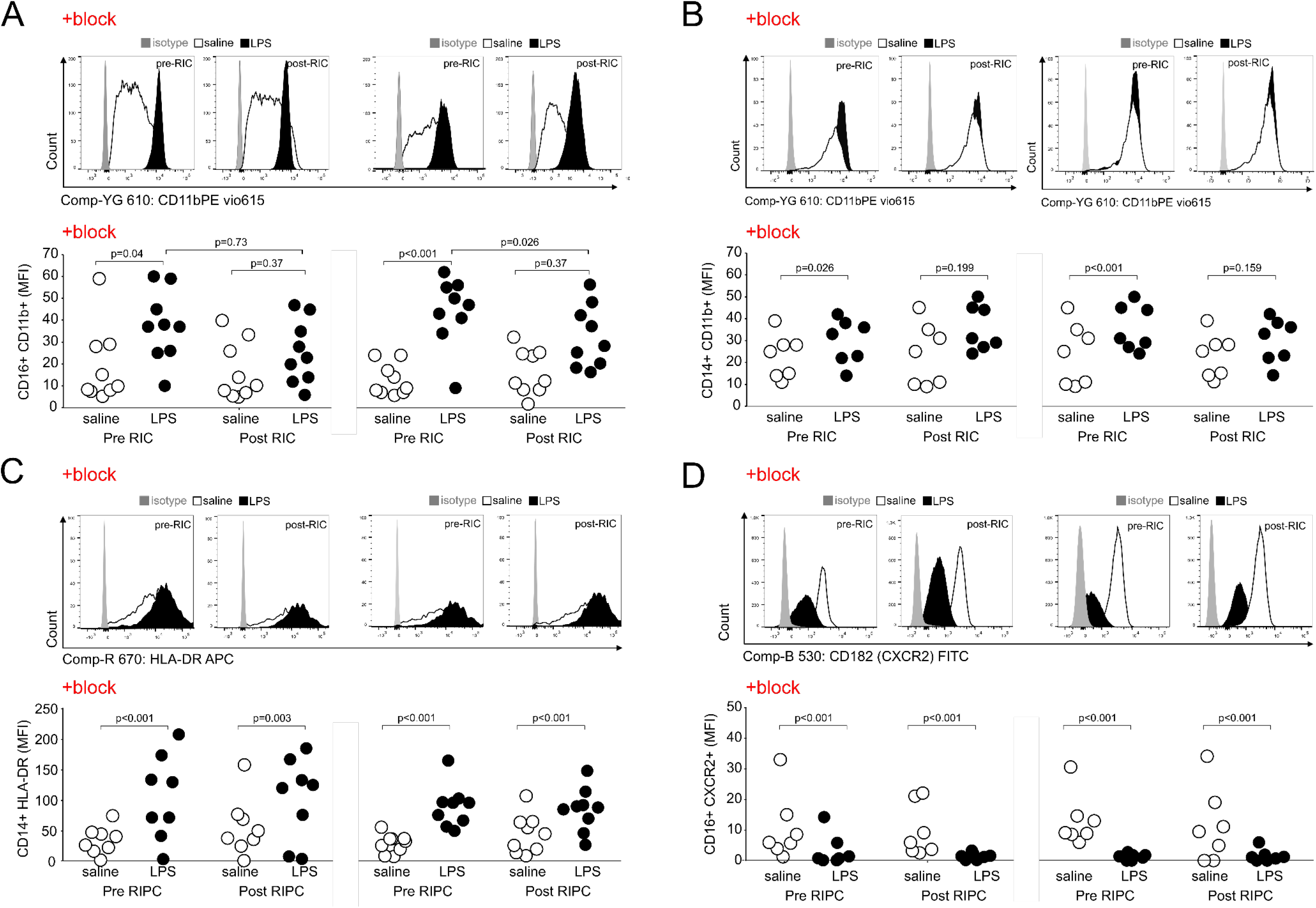
Flow cytometry before and after RIC in seven individuals with/without brachial plexus block. A. Surface expression of CD11b on circulating CD16 neutrophils. B. Surface expression of CD11b on circulating CD14 monocytes. C. Surface expression of HLA-DR on circulating CD14 monocytes. D. Surface expression of CXCR2 (CD182) on circulating CD16 neutrophils. P values are shown for post-hoc Tukey-Kramer testing (RIC/pre RIC versus LPS/saline control).

After establishing brachial plexus blockade, expression of CD11b on neutrophils remained similar (P=0.72; Supplementary figure 3). RIC after establishing brachial plexus blockade attenuated the LPS-induced increase in CD11b expression (mean difference in MFI: 9 (95%CI:-1 to 19; P=0.08; post-hoc Tukey-Kramer test). When subjects returned six weeks later to undergo RIC alone, LPS again increased surface expression (median fluorescence intensity) of CD11b on CD16^+^ neutrophils by 31 units (95%CI:25-38); P<0.001; post-hoc Tukey-Kramer test) (Figure 6A). RIC had no effect on CD16CD11b expression (mean difference:-3 (95%CI:-10 to 3); P=0.41). After RIC, the effect of LPS on surface expression of CD11b on neutrophils was reduced, compared to the effect of LPS before RIC (mean difference:12 arbitrary units (95%CI:5-18); P=0.002; post-hoc Tukey-Kramer test). CXCR2 expression on neutrophils, which mediates massive cardiac neutrophil infiltration after myocardial infarction,(20) was similar between RIC and RIC_block_. CXCR2 expression on neutrophils after incubation with LPS were similar in the presence, or absence, of brachial plexus blockade (Figure 6D).

### Monocyte activation

Before establishing brachial plexus blockade prior to RIC, LPS increased surface expression of CD11b on CD14^+^ monocytes (mean difference:9 (95%CI:1-18); P=0.04; Figure 6B). RIC had no effect on CD14^+^CD11b expression (mean difference:-1 (95%CI:-25 to 23); P=0.98). After RIC, LPS had no effect on surface expression of CD11b on monocytes (mean difference:-7 arbitrary units (95%CI:-30 to 17); P=0.73). When subjects returned six weeks later to undergo RIC alone, LPS increased surface expression of CD11b on monocytes before RIC was implemented (by 17 arbitrary units (95%CI:7-28); P=0.001; post-hoc Tukey-Kramer test; Figure 6A). After RIC, LPS failed to increase surface expression of CD11b (mean difference:-5 (95%CI:-16 to 5); P=0.38; post-hoc Tukey-Kramer test). The presence or absence of brachial plexus block had no effect on LPS-evoked increases in HLA-DR expression on CD14^+^ monocytes (Figure 6C). LPS evoked a similar increase in HLA-DR after RIC (39 (95%CI:27-52); P<0.001), but this response was similar in magnitude to the effect seen before RIC (mean difference:4 arbitrary units (95%CI:-8 to 17); P=0.71).

## Discussion

This study demonstrates that RIC preserves cardiac vagal activity through afferent signalling from the organ/tissue undergoing RIC, as measured by three independent measures of heart rate variability. When neural transmission from ischaemic tissue was prevented by brachial plexus block using local anaesthesia, vagal activity was reduced. Acute inflammation elicited ex-vivo by LPS augmented markers of myeloid activation following brachial plexus block, but this inflammatory response was reduced by RIC when afferent signalling from the ischaemic tissue was intact.

A role for neural afferents in RIC has been suggested in experimental models in which peripheral nociception induced by skin incisions on the abdomen provided cardioprotection in mice.(13) Topical application of 0.1% capsaicin cream on the abdomen or paw before myocardial ischaemia/reperfusion (6) (12) reduced infarct size in rodents via neurogenic signaling involving spinal nerves, sympathetic nerves, and activation of PKCε in the heart. (13) Similarly, chronic neuropathic pain activates neurons within the anterior nucleus of paraventricular thalamus, which increases release of acetylcholine via the vagus nerve activity to trigger PKCε-mediated cardioprotection following ischemia-reperfusion injury.(21) Pharmacological inhibition of extracellular signal-regulated kinase activation in the PVA abolishes neuropathic pain-induced cardioprotection, whereas pharmacologic or optogenetic activation of PVA neurons confers cardioprotection.(21) By extension, patients with established peripheral neuropathy (e.g. those with diabetes mellitus),(22, 23) or who acquire loss of vagal activity (e.g. central neuromodulatory effects of general anaesthesia),(16, 24) are therefore not likely to benefit from RIC.

We used three measures of heart rate variability to dissect the autonomic components contributing to RIC. The 10 minute recording period at the start and end of each protocol enabled sufficient time to capture high quality ECG data for the analysis. Frequency domain and non-linear analyses strongly suggested that parasympathetic activation is a key feature of RIC, being independent of underlying changes in heart rate.(25) We performed spectral analysis using the parametric autoregressive method (AR) because it produces a spectrum with superior resolution using shorter data frames.(26) We used HF as an index of vagal modulation of heart rate, which is abolished under conditions of vagal blockade.(17) RIC alone preserved the high frequency power band whereas HF declined after RIC in the presence of brachial plexus block, suggesting neural afferent activity is required to maintain parasympathetic activity. In support of this, the non-linear measure DFA, which most closely corresponds to short-term fluctuations, increased after RIC following brachial plexus block. Given that DFAα1 was preserved in the absence of brachial plexus block, these data suggest that RIC initiates vagal activation since DFAα1 increases with vagal blockade and decreases with sympathetic blockade.(19)

The rapid effect of RIC in conferring cardioprotection – over minutes-led us to focus primarily on circulating myeloid cells. As the archetypal first responders to acute inflammation and injury, neutrophils are activated rapidly. Neutrophil depletion reduces tissue injury after myocardial ischemia-reperfusion in both patients(27) and animal models of myocardial ischaemia-reperfusion.(28) However, dysregulated, persistent and/or over-exuberant leukocyte recruitment to ischaemic tissue can fuel excessive inflammation and exacerbate further tissue injury.(29) Our findings are the first to provide human data in support of a direct link between preserved efferent vagal activity and neutrophil responses to ischaemic-reperfusion injury. The surface receptor integrin CD11b/CD18 serves as a pattern recognition receptor on phagocytes, including neutrophils, to recognize pathogen and damage-associated molecular patterns.(30) A specific anti-CD18 monoclonal antibody reduces polymorphonuclear cell-mediated contractile dysfunction in an in vitro model of myocardial ischemia-reperfusion injury by limiting polymorphonuclear cell accumulation.(31)

RIC failed to prevent LPS-induced downregulation of CXCR2 and increased expression of monocyte HLA-DR. Our observations are consistent with laboratory models exploring the vagal anti-inflammatory reflex, which demonstrated a reduction, rather than complete suppression, of the inflammatory response to a range of DAMPs and PAMPs.(32) Nicotinic cholinergic receptors provide the neuroimmune receptor mediated link between myeloid cells and preserved vagal activity.(33) Unstimulated neutrophils isolated from human blood express α7nAChRs and α3β4 nAChRs.(34) Nicotine, an agonist of the α7nAChR mediated vagal anti-inflammatory pathway in macrophages, reduces levels of CD11b on the surface of neutrophils in a dose-dependent manner by suppressing F-actin polymerization, the rate-limiting step for CD11b surface expression.(3) As with macrophages, vagus nerve stimulation attenuates neutrophil surface CD11b levels only in the presence of an intact and innervated spleen.(3) Reduced expression of CD11b, which also serves as a receptor for complement C3b, limits cell-mediated cytotoxicity, chemotaxis and phagocytosis.

A rapid decline in the surface expression of HLA-DR is observed on monocytes obtained from patients after acute myocardial infarction.(35) Circulating CD14^+^HLA-DR^neg/low^ monocytes secrete high levels of TNFα, IL-6, and IL-1ß which promote proinflammatory immune responses; in the expansion of this monocyte population after myocardial infarction correlates with both cardiac damage and physiological function.(36) However, RIC had no effect on CD14^+^HLA-DR surface expression either in the presence, or absence, of brachial plexus block. Similarly, expression of the chemokine receptor CXCR2, deficiency of which limits neutrophil recruitment and the extent of myocardial infarction size, was not altered by RIC in the presence or absence of brachial plexus block.

Several randomised clinical trials, particularly in elective cardiac surgery, have failed to show consistent benefits of RIC on clinical outcomes.(37) Our study provides direct human translational data supporting laboratory findings demonstrating that autonomic modulation contributes to the downstream biologic effects of RIC. Bidirectional feedback mechanisms between the heart and the brain require both neural and humoral pathways for effective RIC.(38) The frequency and ease with which the autonomic component of RIC may be disrupted is likely to account for the results in several neutral trials. For example, peripheral neuropathy in patients with diabetes mellitus and metabolic syndrome is common.(39) Markedly reduced vagal activity is common in individuals who are physically deconditioned.(40-42) Furthermore, neural processing in autonomic pathways within the central nervous system is profoundly disrupted by anaesthetic agents.(43, 44)

The development of this human model enabled each subject to serve as their own control while remaining masked to laboratory-based outcomes. Capturing both autonomic and immunologic readouts added further complementary data to address the central hypothesis that RIC preserves and/or stimulates vagal activity RIC. We were unable to randomise the sequence of block versus control RIC, as subjects gave consent prior to elective upper limb surgery. Similarity in both autonomic and inflammatory measures at the start of both control and block protocols suggest that the impact of randomising the sequence of interventions would not be significant. Brachial plexus nerve block is not a minor procedure for subjects who do not require surgery, hence the choice of participants were preoperative patients. It is important to highlight that ultrasound-guided brachial plexus block was undertaken without sedation, which alters autonomic function.(45) Although data were captured only during the protocol period, we have established that this model of brachial plexus block serves as a potentially useful tool over the longer term to further dissect mechanisms of RIC in humans.

In summary, we have shown, for the first time in humans, that RIC requires an afferent neural component to preserve vagal and recruit anti-inflammatory mechanisms previously identified in laboratory animal models. Closing this translational gap in mechanistic knowledge in humans explains, in part, the variable effect of RIC in conferring cardiac and renal protection.(38) Our data suggest that a personalised medicine approach may benefit individuals who are capable of mounting an integrated innate protective reflexes in response to RIC.

## Data Availability

Data availability as referred to in the manuscript: requests to corresponding author.

## Notes

### Competing Interest Statement

The authors have declared no competing interest.

### Clinical Trial

researchregistry6482

### Funding Statement

This work was supported by the British Heart Foundation (Ref: RG/14/4/30736 and RG/19/5/34463); British Journal of Anaesthesia/Royal College of Anaesthetists Basic Science Career Development Award; British Oxygen Company research chair grant, Royal College of Anaesthetists; UK National Institute for Health Research.

### Author Declarations

London (Stanmore) Research Ethics Committee (16/LO/0634).

