## Supplementary data for "Activation of the vagal anti-inflammatory reflex by remote ischaemic conditioning in humans: experimental cross-over study"

### Contents

Supplementary Figure 1. Gating strategy ........ 2
Supplementary Figure 2. Time domain measures of HRV. ........ 3
Supplementary Figure 3. Lack of acute effect of brachial plexus block on flow cytometry measures. . 4

### Supplementary Figure 1. Gating strategy

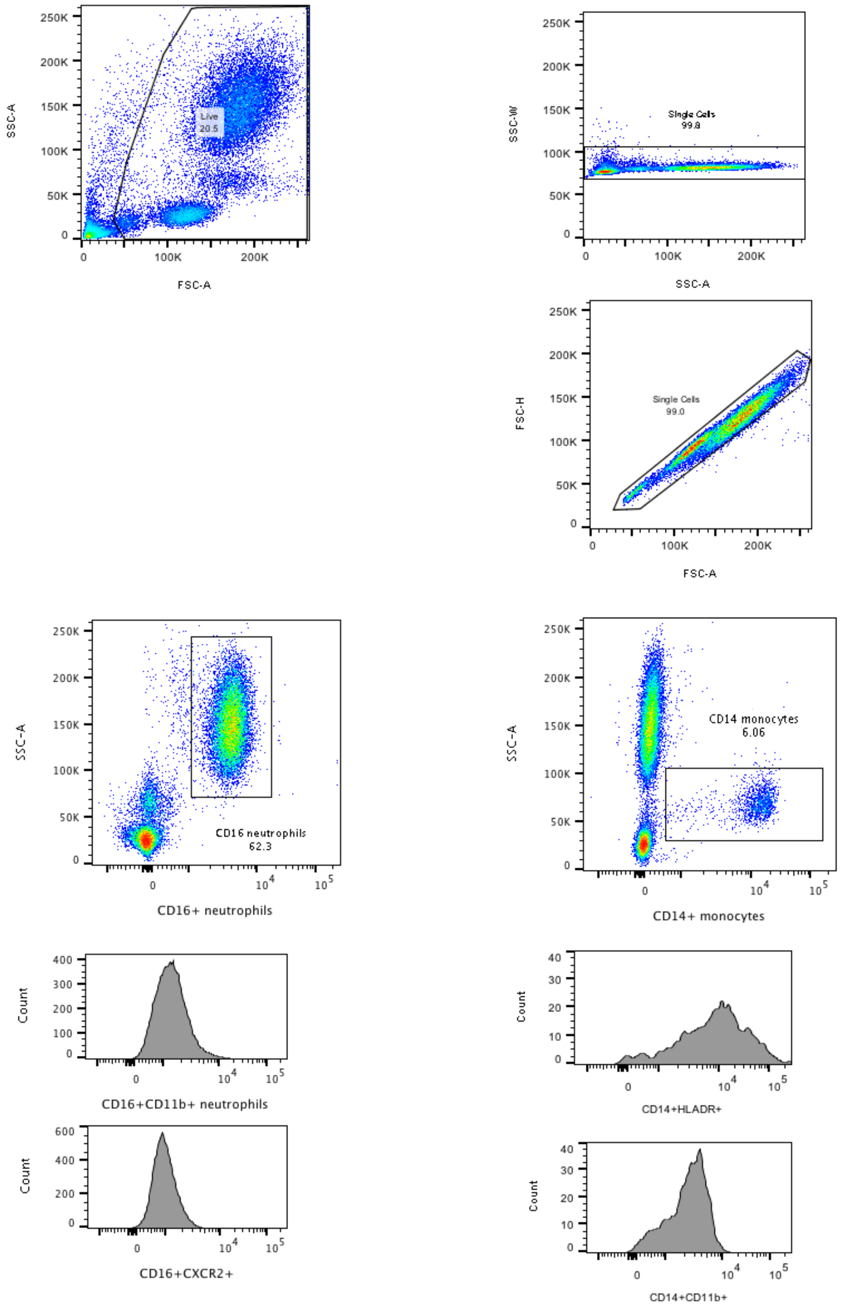

**Supplementary Figure 2. Time domain measures of HRV.**

A. SDNN, the standard deviation of NN intervals
B. RMSSD, Root Mean Square of the Successive Differences in RR interval.

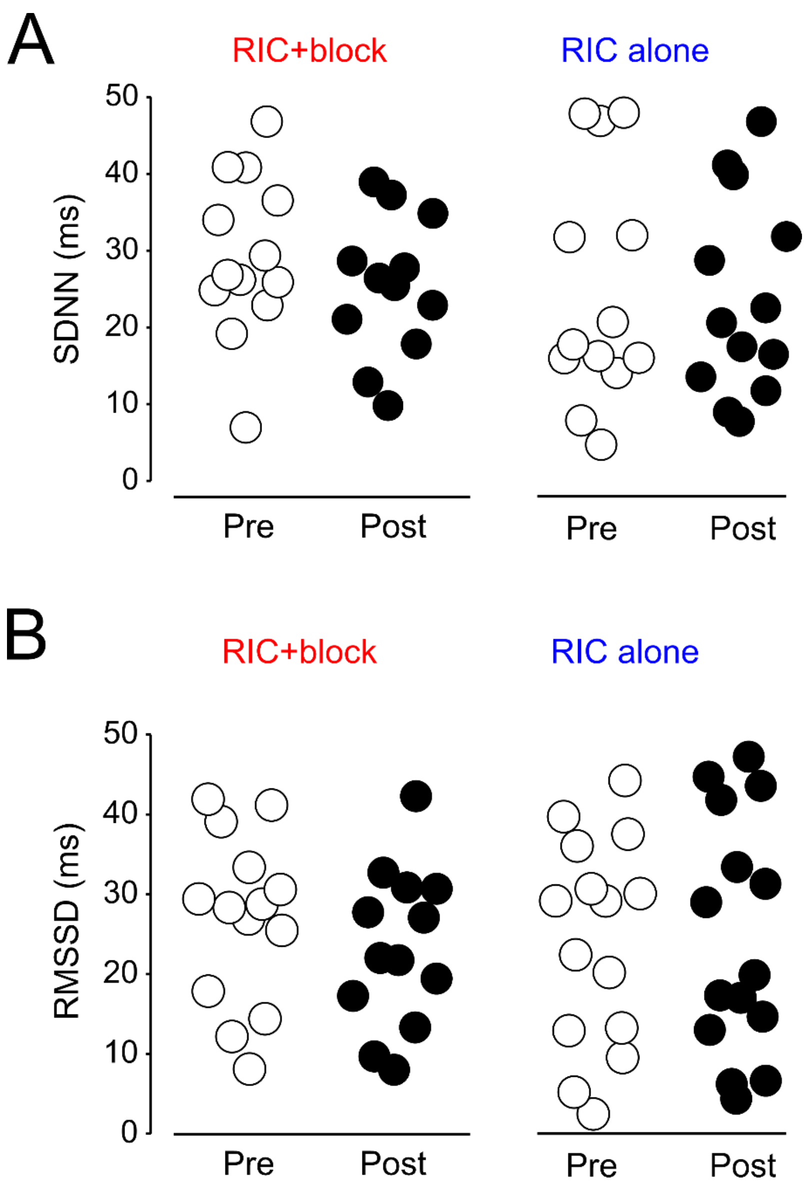

**Supplementary Figure 3. Lack of acute effect of brachial plexus block on flow cytometry measures.**

Representative plots from same individual before and after injection of local anaesthetic into brachial plexus.

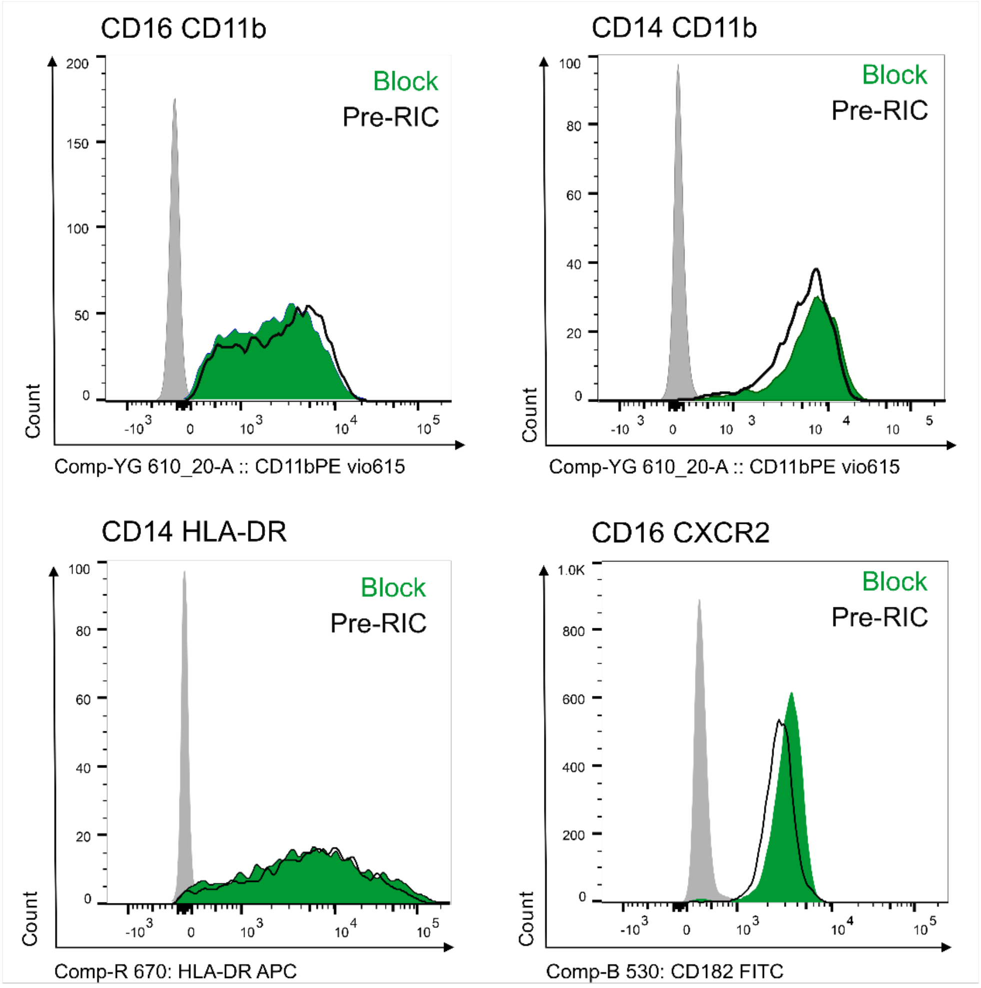
